# Mechanisms and kinetics of bacterial clearance after experimental colonisation in adults with asthma

**DOI:** 10.1101/2020.08.19.20177790

**Authors:** Seher Raza Zaidi, Simon P. Jochems, Jesús Reiné, Sherin Pojar, Elissavet Nikolaou, Elena Mitsi, Esther L. German, Angela D. Hyder-Wright, Hugh Adler, Helen Hill, Caroline Hales, Victoria Connor, Carla Solórzano, Stephen B. Gordon, John Blakey, David Goldblatt, Daniela M. Ferreira, Jamie Rylance

## Abstract

**Background:** Pneumococcal pneumonia is a leading cause of death, particularly affecting those with chronic respiratory disease. Observational studies suggest increased nasopharyngeal colonisation rates with *S.pneumoniae* in asthma, and lower specific antibody levels.

**Objectives:** Using experimental human pneumococcal challenge, we examined the acquisition and kinetics of nasopharyngeal colonisation of *Streptococcus pneumoniae* serotype 6B. We also aimed to dissect associated mucosal and systemic immune responses and immunizing effect of carriage.

**Methods:** Fifty participants with asthma well-controlled on moderate inhaled corticosteroid doses were challenged with pneumococcus, and a subset of colonized individuals were re-challenged 6–11 months later with the same pneumococcal isolate. Colonisation rates (from nasal wash), systemic antibody levels and mucosal cellular and cytokine responses were compared to 151 healthy controls.

**Measurements and Main Results:** Colonisation rates were 28/50 (56%) and 68/151 (45%) in those with asthma and controls respectively, p=0.17. Duration of colonisation was shorter in people with asthma (median 14 days vs 29 days, p=0.03) but of similar density. Body mass index was higher in colonised compared with non-colonised asthma individuals (median 24.7 [IQR 24.1–29.0] and 23.5 [20.1–26.4] respectively, p=0.019). Despite an increase in pneumococcal capsular and protein antibodies after colonisation, 4/12 asthmatic individuals became colonised again upon re-challenge. Nasal neutrophil and T cell levels, in particular mucosa associated invariant T (MAIT) cells were decreased in people with asthma compared to healthy controls (median 9.4, [IQR 5.0–13.3 %] of CD8^+^ T cells) vs median 15.8, [IQR 9.9–25.9 %] of CD8^+^ T cells respectively (p=0.0047). Most nasal cytokines were also reduced in asthmatics. In both groups, colonisation led to recruitment of monocytes and granulocytes to the nasal mucosa.

**Conclusions:** Nasopharyngeal colonisation was of shorter duration in those with asthma compared to controls, although acquisition rates were not different. Rates of colonisation were higher with increasing BMI in individuals with asthma. Despite a baseline reduction in mucosal immune cells and cytokines in asthmatics with corticosteroids, colonisation led to cellular recruitment in both groups. Colonisation was not associated with protection from homologous re-challenge in individuals with asthma, in contrast to healthy volunteers.

**Clinical Implication: (single sentence):** People with asthma on inhaled corticosteroids have an increased likelihood of pneumococcal infection secondary to reduced mucosal immune responses from nasopharyngeal colonisation and a lack of protection from re-exposure.

**Capsule Summary:** Epidemiological studies show that people with asthma are more likely to have nasal colonisation with *S. pneumoniae*, which may proceed to infection such as pneumonia and invasive pneumococcal disease. This study investigates the mechanisms underlying pneumococcal colonisaion and its effect on subsequent pneumococcal encounters.

## Introduction

*Streptococcus pneumoniae* is the most common cause of bacterial pneumonia. Large scale real-world observational data shows people with asthma on inhaled corticosteroids (ICS) are at increased risk of pneumonia in a dose-dependent manner ^1–3^, though this relationship was not observed in manufacturers’ retrospective analysis of randomised clinical trials. Epidemiologically, asthma is associated with an increased prevalence of nasopharyngeal colonisation by *S. pneumoniae*^4– 7^, although information on density and duration are lacking. An Italian cross-sectional study reported a 45% colonisation rate in children and adolescents with asthma ^4^ and nasopharyngeal swabs from Finnish army recruits identified those with asthma twice likely to be colonised with *S.pneumoniae^8^*. Mucosal cellular mechanisms are important for clearance of nasopharyngeal colonisation of *S. pneumoniae ^9^*, and an impairment of these may lead to increased colonisation and perhaps acute infection such as pneumonia and invasive pneumococcal disease^10^. Airway inflammation in asthma, and ICS therapy, alter mucosal immune responses ^11–13^. Routine immunisation against pneumococcus is offered to people with chronic respiratory disease and those deemed immune compromised secondary to treatment with oral corticosteroids in the United Kingdom and United States ^1, 4, 5^.

We hypothesise that the immunological containment of pneumococcal colonisation is altered in asthma due to the condition itself, or secondary to ICS and that altered mucosal dynamics of colonisation mediate susceptibility to disease. We used an established human challenge model ^14, 15^ to determine factors underlying susceptibility to pneumococcal colonisation in people with asthma and healthy controls^16, 17^.

## Methods

### Study Design

We recruited 50 participants, from June 2016 until April 2018, aged 18–50 years with physician-diagnosed asthma. All were receiving ICS therapy at British Thoracic Society (BTS) treatment steps 2 (up to 400µg budesonide equivalent [BDP] per day) or step 3 (up to 800μg BDP per day and additional long acting beta agonist [LABA]) – see Table 1 and Table E1. All participants gave informed consent.

**Table 1:**
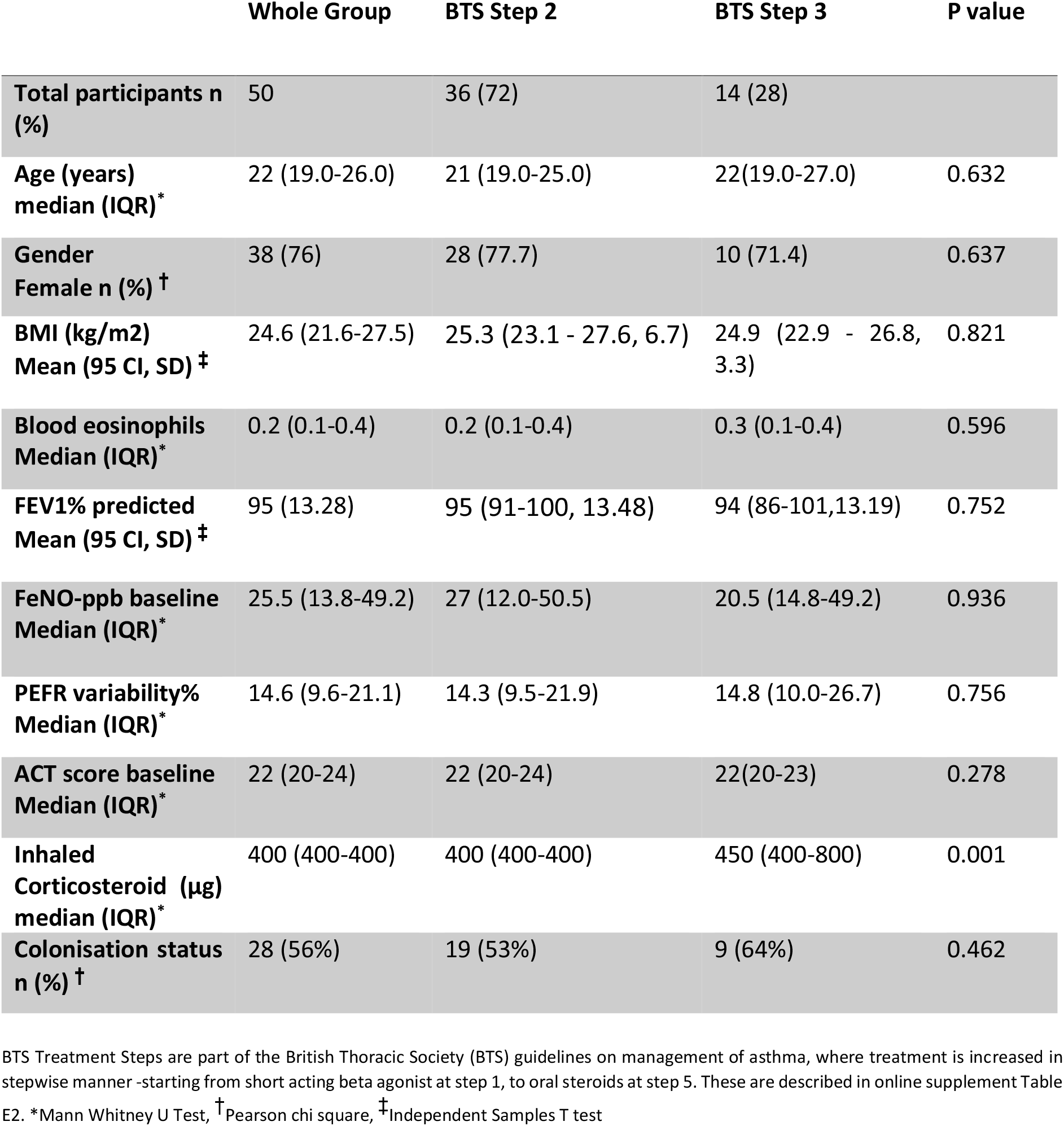
Demographics and clinical characteristics for the asthma cohort according to BTS treatment step

The participant demographics and baseline characteristics were described by clinical history, spirometry with reversibility by inhaled β_2_ agonist according to ATS guidelines (EasyOne®, ndd Medical Technologies, Switzerland), fractional exhaled nitric oxide (Niox Vero®, Circassia, UK), and blood eosinophil count. Information was sought from primary care to confirm the medical history, current treatment and allergies. At screening, all participants were provided a written asthma action plan, with a peak expiratory flow rate meter and diary (Mini Wright®, Clement and Clarke Limited, UK). Exclusion criteria included: forced expiratory volume less than 70% predicted after salbutamol; more than 1 exacerbation in the preceding year; treatment with antibiotics or oral steroids in the preceding 4 weeks (full list of exclusion criteria in Table E2). Inoculation was performed as previously described ^14,18^, by instillation of 80,000 colony-forming units (CFU) *Streptococcus pneumoniae* serotype 6B in 100µL saline into each nostril. This strain has been previously used for challenge by us and others with reproducible carriage rates, is fully sequenced, amoxicillin-sensitive, and does not cause any natural colonisation in Liverpool ^14, 15, 18^. Follow-up visit schedules are described in Figure E1, and nasal sampling was performed as described for previous studies^14, 15, 19^.

Participants were contacted daily for the first 7 days to report their peak flow and temperature. They maintained a peak flow diary and symptom log and were advised to follow their personal asthma action plan. At study completion, a 3 day-course of amoxycillin was prescribed for participants who remained colonised at either of the last 2 visits.

For microbiological and immunological comparison we used age-matched healthy controls from an influenza vaccine study (Sep 2015- Mar 2017), with identical methods and sampling, except that influenza-study participants received intramuscular tetravalent inactivated influenza vaccine either 3 days before or after inoculation with pneumococcus ^20^. A subset of colonised asthmatic individuals (n=12) were re-challenged following the same protocol 6–11 months after the initial inoculation event.

### Ethical approvals

Ethical Approval was obtained from the Liverpool East NHS Research Ethics Committee (reference number NW/016/0124). The study was co-sponsored by the Royal Liverpool University Hospital and the Liverpool School of Tropical Medicine. International Randomised Controlled Trial Number (ISRCTN) 16755478). Control group was obtained from the study approved under Liverpool East NHS Research Ethics Committee (EudraCT 2014–004634–26)

### Flow cytometry analysis

#### Nasal

Nasal cells were collected by scraping the inferior turbinate of volunteers consecutively with two curettes (Rhino-pro®, Arlington Scientific), as described before ^19^. Curettes were collected into PBS + 5mM EDTA and 0.5% heat inactivated FBS. Cells were dislodged from curettes by pipetting up and down and immunophenotyping of nasal cells was performed as previously described ^9, 19^. Briefly, cellular samples were stained on ice for 15 minutes with LIVE/DEAD® Fixable Violet Dead Cell Stain (Thermofisher®), followed by staining for 15 minutes on ice with an antibody cocktail containing Epcam-PE, HLADR-PECy7, CD66b-FITC (all Biolegend®), CD3-APCH7 (BD Biosciences®) or CD3-APCCy7 (Biolegend®), CD14-PercpCy5.5 (BD Biosciences®) and CD45-PACOrange (ThermoFisher®). Some samples were additionally stained concurrently with BDCA2-APC (Miltenyi®) and/or CD16-APC, CD1c-PE/Dazzle594, CD19-BV650, CD4-BV605, CD8-BV785, TCRvα7.2-PE/Dazzle594, TCRvα7.2-BV711, CD123– BV711 and/or CD117–BV650 (all Biolegend®). Cells were washed at the end of incubation with 3mL PBS and filtered through a 70µm mesh (ThermoFisher®). After centrifugation for 5 minutes at 450xg, pellets were resuspended in 200µL CellFIX™ (BD Biosciences®) and kept on ice until sample acquisition. Flow cytometry data were acquired on a LSRII cytometer (BD), and analysed using Flowjo X (Treestar®). Compensation matrices were set using compensation beads (Thermofisher^®^). All antibodies were titrated and fluorescence minus one controls were used to verify specificity of signal for each of the antibodies included for at least two samples. Samples with fewer than 500 immune cells or 250 epithelial cells were excluded from analysis. To adjust for variable amount of cells collected between curettages, we analysed both absolute cell counts and cell counts normalized to number of epithelial cells. This has the advantage that an increase or decrease of certain immune populations does not affect frequencies of other immune populations.

#### Peripheral Blood

To stain cells, PBMCs were thawed with 50 μg/mL DNAse I (MilliporeSigma) in prewarmed RPMI containing 10% FBS and washed once in media including DNAse I. Briefly, cellular samples were stained for 15 minutes with LIVE/DEAD® Fixable Violet Dead Cell Stain (Thermofisher®), followed by staining for 15 minutes with an antibody cocktail containing CD38-FITC, CD7-PE, CD196-PEeFluor610, CD5-PECy7 (all from ThermoFisher®), CD161-APC (BD Biosciences®), CD4-PerPCy5.5, CD3-APCCy7, CD8-AlexaFluor700, CD45-BV510, CD69-BV650 and TCRVα7.2-BV786 (all from Biolegend®). Finally, cells were washed, resuspended in 200µL PBS, and acquired. Flow cytometry data were acquired on a LSRII cytometer (BD), and analysed using Flowjo X (Treestar®). Compensation matrices were set using compensation beads (Thermofisher ®).

### Mucosal cytokine levels

Concentrated nasal lining fluid was collected by applying synthetic absorptive matrices to the nasal mucosa for 1 minute and stored at –80°C until analysis ^21^. Fluid was eluted by adding 100µL Luminex assay buffer, supernatant was clarified by centrifugation for 10 minutes, and 30 cytokines and immune proteins were analysed as per manufacturer’s protocol as we previously described ^9, 19^.

### Antibody titres

Titres of antibodies against pneumococcal capsule and proteins were measured as previously described ^15^. Briefly, ELISA plates were coated with 6B polysaccharide at 4°C overnight and washed 3 times. Absorption buffer was used for blanks, and to generate standards through serial dilution of 89SF. Samples were diluted 1 in 100, and assayed in duplicate. After loading, plates were incubated at room temperature for 2 hours, washed 3 times in washing buffer, and secondary antibody applied and incubated for 90 minutes. Following 3 further washes, plates were incubated in the dark with 100μl/well of para-nitrophenyl phosphate, and absorbance measured at 405nm. Samples with coefficient of variation>15% between duplicates were repeated and one sample due to a very high signal (outside standard curve) was repeated using a 1 in 800 dilutions.

#### MSD

In a WHO reference lab, ELISA plates were coated with proteins by MSD. Standards were prepared using 007sp in an initial 1:100 dilution with subsequent four-fold serial dilution. Samples were diluted 1:100, and assayed in duplicate with quality control (QC) sera at 1:1000 and 1:2000 dilutions. Reactions were stopped by agitating with 150μL/well blocking solution (5% BSA 1×PBS) at 700rpm for 1 hour at room temperature. After 4 washes, detection antibody was added for a further 1 hour on the shaker, before a final wash and immediate reading.on a MSD imager.

### Statistical Analysis

SPSS version 22, R 3.5.0 and Graphpad prism version 5 were used for analysis. Area under the bacterial density-time curve were calculated by trapezoid method of log_10_ (value+0.01). Colonisation rates were compared by chi-square test, and continuous measures by Mann-Whitney U test and independent samples T-test for non-normal distributed and normal distributed data, respectively. Spearman and Wilcoxon tests were used for correlation and paired-sample analysis respectively. All tests were two-tailed, with significance level of p< 0.05.

### Primary End Point and Sample Size Calculation

The primary end-point was the experimental colonisation rate in asthmatics compared to healthy controls. Experimental colonisation was defined as positive bacterial culture from nasal wash at any point after inoculation until the end of follow-up. A sample size of 52 was estimated from historic challenge study data in healthy volunteers (colonisation rate 49.6%) and assuming 50% reduction in colonisation rate in people with asthma (i.e. absolute risk difference 24.6%).

## Results

A total of 95 volunteers were consented, of which 50 were subsequently inoculated with *Streptococcus pneumoniae* 6B (Figure E2). Reasons for exclusion are described in detail in Figure E2, the commonest being lack of ICS therapy. Median age in the asthma and control groups was 22 years [IQR 20–26] and 20 years [IQR 19–22] respectively (Table 3). 38 (76%) asthma participants and 82 (54%) controls were female (Table 1–3). No serious adverse events occurred. Median baseline blood eosinophil count was 0.2 [IQR 0.1–0.4], with median FEV1% predicted 95% [91–100], median FeNO 25.5 [13.8–49.2] and asthma control (ACT) score 22 [20–24] (ACT 25 point scale – score 20–24 well controlled, less than 20 not well controlled).

**Table 3.**
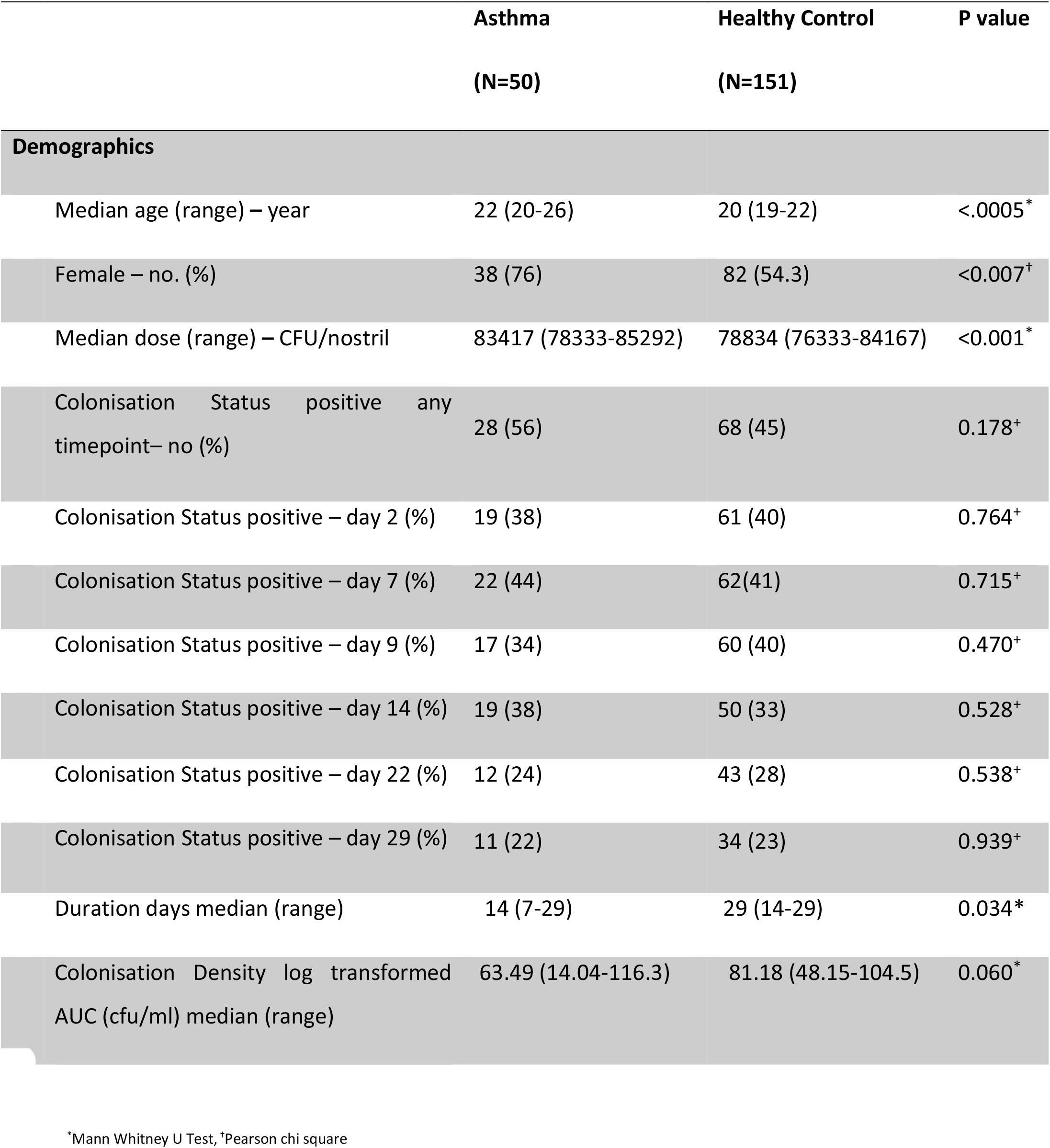
Colonisation rates in people with asthma compared to healthy control

|  | Asthma<br>(N=50) | Healthy Control<br>(N=151) | P value |
| --- | --- | --- | --- |
| **Demographics** |  |  |  |
| Median age (range) – year | 22 (20-26) | 20 (19-22) | <.0005* |
| Female – no. (%) | 38 (76) | 82 (54.3) | <0.007<sup>†</sup> |
| Median dose (range) – CFU/nostril | 83417 (78333-85292) | 78834 (76333-84167) | <0.001* |
| Colonisation Status positive any<br>timepoint– no (%) | 28 (56) | 68 (45) | 0.178<sup>+</sup> |
| Colonisation Status positive – day 2 (%) | 19 (38) | 61 (40) | 0.764<sup>+</sup> |
| Colonisation Status positive – day 7 (%) | 22 (44) | 62(41) | 0.715<sup>+</sup> |
| Colonisation Status positive – day 9 (%) | 17 (34) | 60 (40) | 0.470<sup>+</sup> |
| Colonisation Status positive – day 14 (%) | 19 (38) | 50 (33) | 0.528<sup>+</sup> |
| Colonisation Status positive – day 22 (%) | 12 (24) | 43 (28) | 0.538<sup>+</sup> |
| Colonisation Status positive – day 29 (%) | 11 (22) | 34 (23) | 0.939<sup>+</sup> |
| Duration days median (range) | 14 (7-29) | 29 (14-29) | 0.034* |
| Colonisation Density log transformed<br>AUC (cfu/ml) median (range) | 63.49 (14.04-116.3) | 81.18 (48.15-104.5) | 0.060* |

\*Mann Whitney U Test, <sup>†</sup>Pearson chi square

Comparing individuals receiving step 2 and 3 treatment, there was no difference in blood eosinophils (p=0.596) and FeNO (p = 0.936), or control as measured by Asthma Control Test (p=0.278) (Table 1). There was a positive correlation between FeNO levels and blood eosinophil levels (p<0.0001, r=0.573).

### Determinants of colonisation in asthma

Participants who were colonised (positive bacterial culture from nasal wash at any time point) and non-colonised (negative bacterial culture from nasal wash at all time points) had similar baseline characteristics (Table 2). One participant was colonised at baseline with *S. pneumoniae* serotype 9, and subsequently became experimentally colonised with 6B, with both serotypes identified at one time point. Body mass index (BMI) was higher in colonised than in non-colonised participants (median 24.7 [IQR 24.1–29.0] and 23.5 [20.1–26.4] respectively, p=0.019). FeNO levels were not significantly different at baseline (colonised participants median 20.5 [IQR 13.5–37.5] vs non-colonised participants 29.5 [IQR13.5–54.75], p=0.287).

**Table 2.** Demographics and clinical characteristics for the asthma cohort according to colonisation status

|  | Whole Group | Colonised | Non-Colonised | p value |
| --- | --- | --- | --- | --- |
| **Number of Participants** | 50 | 28 (56) | 22 (44) | n/a |
| **Age (years) median (IQR) \*** | 22 (19.8-26.0) | 22 (20-26) | 21(19-24) | 0.432 |
| **Gender Female n (%)<sup>†</sup>** | 38 (76) | 23(82) | 15(68) | 0.251 |
| **BMI (kg/m2) Median (IQR) <sup>‡</sup>** | 24.6 (21.6-27.5) | 24.7 (24.1-29.0) | 23.5 (20.1-26.4) | 0.019 |
| **Blood eosinophils Median (IQR) \*** | 0.2 (0.1-0.4) | 0.2 (0.1-0.45) | 0.25 (0.1-0.4) | 0.933 |
| **<sup>§</sup>Blood Eosinophils >0.3 n (%)<sup>†</sup>** | n/a | 16 (59%) | 11 (41%) | 0.615 |
| **FEV1% predicted Median (IQR) <sup>‡</sup>** | 95.5 (87.5-105) | 98.5 (88.25-105) | 94.5 (84.5-103.5) | 0.340 |
| **PEFR variability >12% <sup>‡</sup>** |  | 15 (62.5%) | 13 (72%) | 0.508 |
| **FeNO-ppb baseline Median (IQR) \*** | 25.5 (13.75-49.25) | 20.5 (13.5-37.5) | 29.5 (13.5-54.75) | 0.287 |
| **FeNO ppb Day 7 median (IQR) \*** | 25 (11-41.5) | 25 (10-37) | 26.5 (13.75- 54.75) | 0.526 |
| **FeNO >40 ppb n \*\* (%)<sup>†</sup>** | n/a | 6 (37.5%) | 10 (62.5%) | 0.071 |
| **ACT score baseline Median (IQR) <sup>‡</sup>** | 22 (20.75-24) | 22 (20.25-24) | 22 (20.75-24) | 0.129 |
| **ACT score Day 29 median (IQR) <sup>‡</sup>** | 22 (21-24) | 22 (22-24) | 22.5 (22-24) | 0.816 |
| **Inhaled Corticosteroid (µg) median (IQR) \*** | 400 (400-400) | 400 (400-500) | 400 (350-400) | 0.208 |

\* Mann Whitney U Test, <sup>†</sup> Pearson chi square, <sup>‡</sup> Independent Samples T test, <sup>§</sup> missing values
<sup>\*\*</sup>n = number of participants with a raised FeNO in each cohort. <sup>||</sup> Pearson's chi square p=0.071 for a raised FeNO of >40 at baseline for colonisation and for change in FeNO from baseline <sup>†</sup> Pearson's chi square p=0.367.
<sup>§</sup>Pearson's Chi square p=0.615 for a raised eosinophil count of >0.3 at baseline, Pearson's chi square test p=0.274 for colonisation positive with steroid dose >400µg, and for steroid dose >500µg p=0.288

### Experimental pneumococcal colonisation in asthmatics vs healthy controls

The median inoculation dose per nostril was 78,834 CFU [IQR 76,333–84,167] for healthy controls compared to people with asthma 83417 CFU [78,333–85,292]. Table 3 for demographics.

Experimental colonisation rates were not significantly different in people with asthma compared with healthy controls (28/50 [56%] vs.68/151 [45%] respectively, p = 0.17). The pneumococcal density as calculated by the area under the density-time curve was also similar in the two groups (Figure 1AµE). However, the duration of colonisation was decreased in asthmatics compared to healthy controls (median 14 days vs 29 days, p = 0.034) (Figure 1F). The area under the density-time curves were similar in asthmatics regardless of the dose of ICS (>400 µg BDP) and BMI>25) (Figure 2A–D).

**Figure 1:**
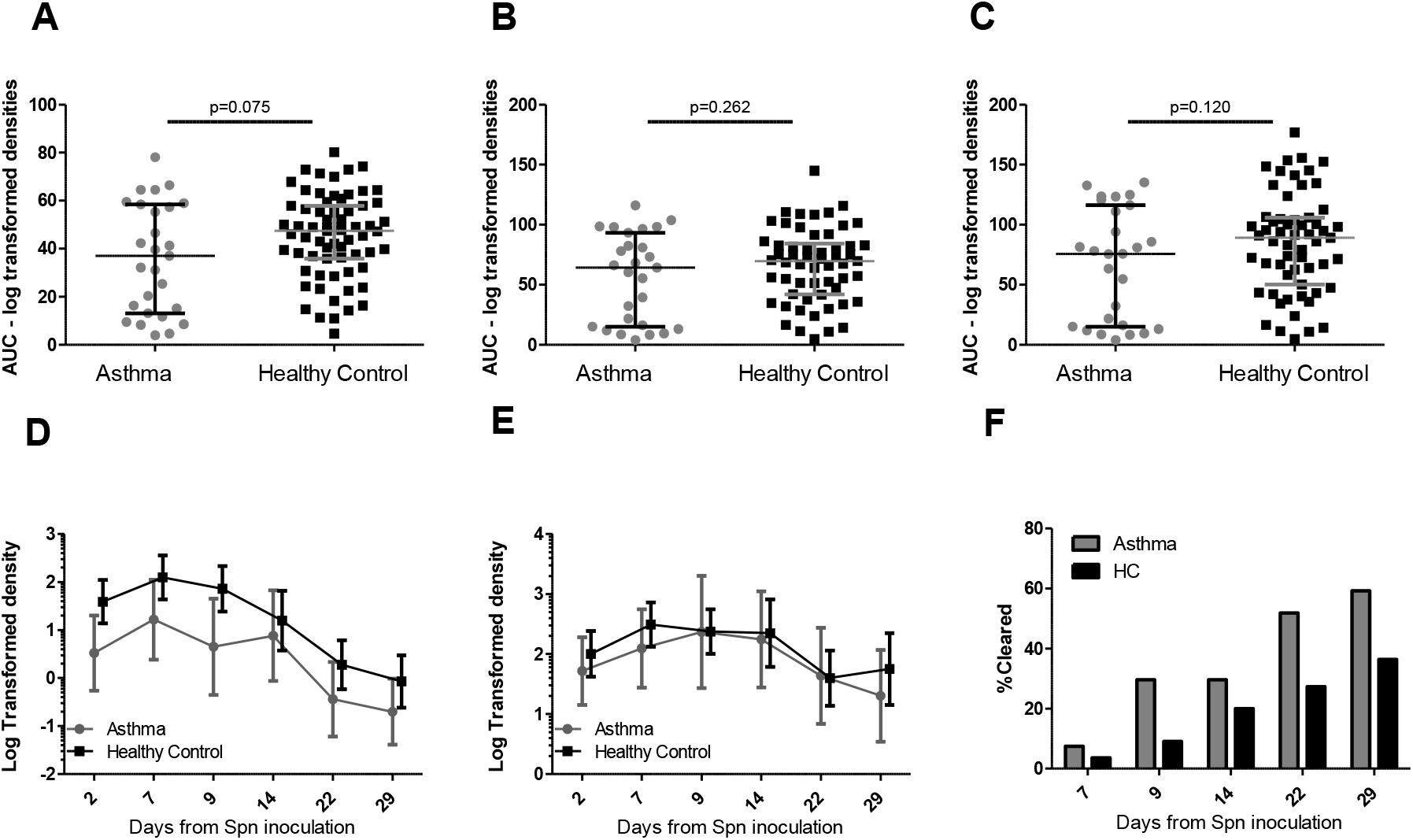
Pneumococcal densities in people with asthma and healthy controls A-C. Area under log-transformed bacterial colonisation density curve for participants colonised (positive bacterial culture from nasal wash) with pneumococcus at any time point who attended all visits up to A) day 14 (n = asthma 27, healthy control (HC) 67), B) up to day 22 (n = 27 asthma, 57 HC) and C) up to day 29 (n = 27 asthma, 59 HC). Individual volunteers are shown, and the lines represent median and inter-quartile range p values for Mann Whitney U Test are shown. D-E: Bacterial colonisation density at each time point; D) all positive participants (positive bacterial culture from nasal wash at any time point) with negative time points included (participants attended the visit and the nasal wash sample was negative for bacterial culture) (n = 28 asthma, 68 HC), E) all positive participants at any timepoint with negative values removed (asthma n=19, 22, 17,19, 12,11 at days 2, 7, 9, 14, 22,29 respectively, HC n=61, 62, 60, 50, 43, 35 at days 2, 7, 9, 14, 22 and 29. Values are mean and 95% confidence interval for log transformed densities at each time point + 0.01 added to all values to allow log transformation). F) Cumulative clearance of colonisation (first time point when nasal wash was negative for bacterial culture following a positive result at an earlier time point) in asthma vs HC at each time point. Clearance rates at each time point for asthma and HC

**Figure 2:**
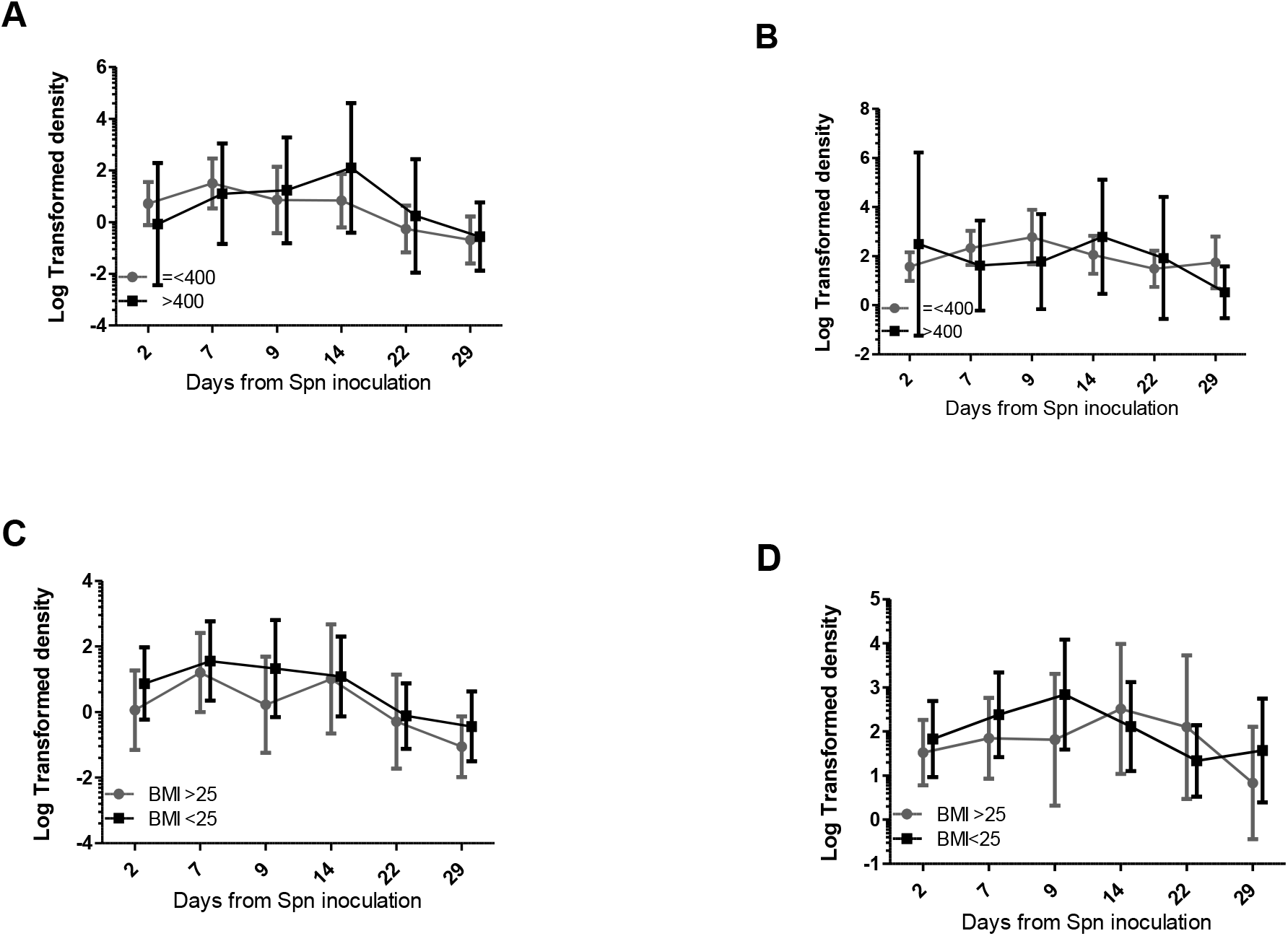
Bacterial colonisation density for asthmatics. A) dose of inhaled corticosteroid (ICS) > 400µg n = 7 or = < 400µg n = 21, all colonised (nasal wash positive for bacterial culture at any time point) participants with negative time points (nasal wash negative for bacterial culture at a specific timepoint) included. B) ICS dose > 400µg or < 400µg all colonised participants with negative time points removed. C) stratification by body mass index (BMI) > 25 n = 12, < 25 n = 16, all colonised participants with negative time points included. D) BMI > 25 or < 25, all colonised participants with negative time points removed. Values are log transformed, bars represent mean and 95% confidence interval.

### Homologous rechallenge of asthmatic individuals

We have reported complete protection in healthy controls against recolonisation with the same 6B strain up to 1 year after initial experimental colonisation (n = 10)^15^. To test this in asthma, twelve colonised individuals were re-challenged with the same strain 6–11 months after the first inoculation. Of these, four became recolonised (33.3%), which is not-significantly different from the 56% colonisation rate during the primary challenge (relative risk 59.5%, 95% CI: 25.8–138%, p = 0.20 with Fisher Exact test).

Within the limitation of our sample size, the density of the second colonisation episode did not appear substantially different from the first (Figure 3A). Of these four individuals, three cleared colonisation within 14 days of the re-challenge.

**Figure 3:**
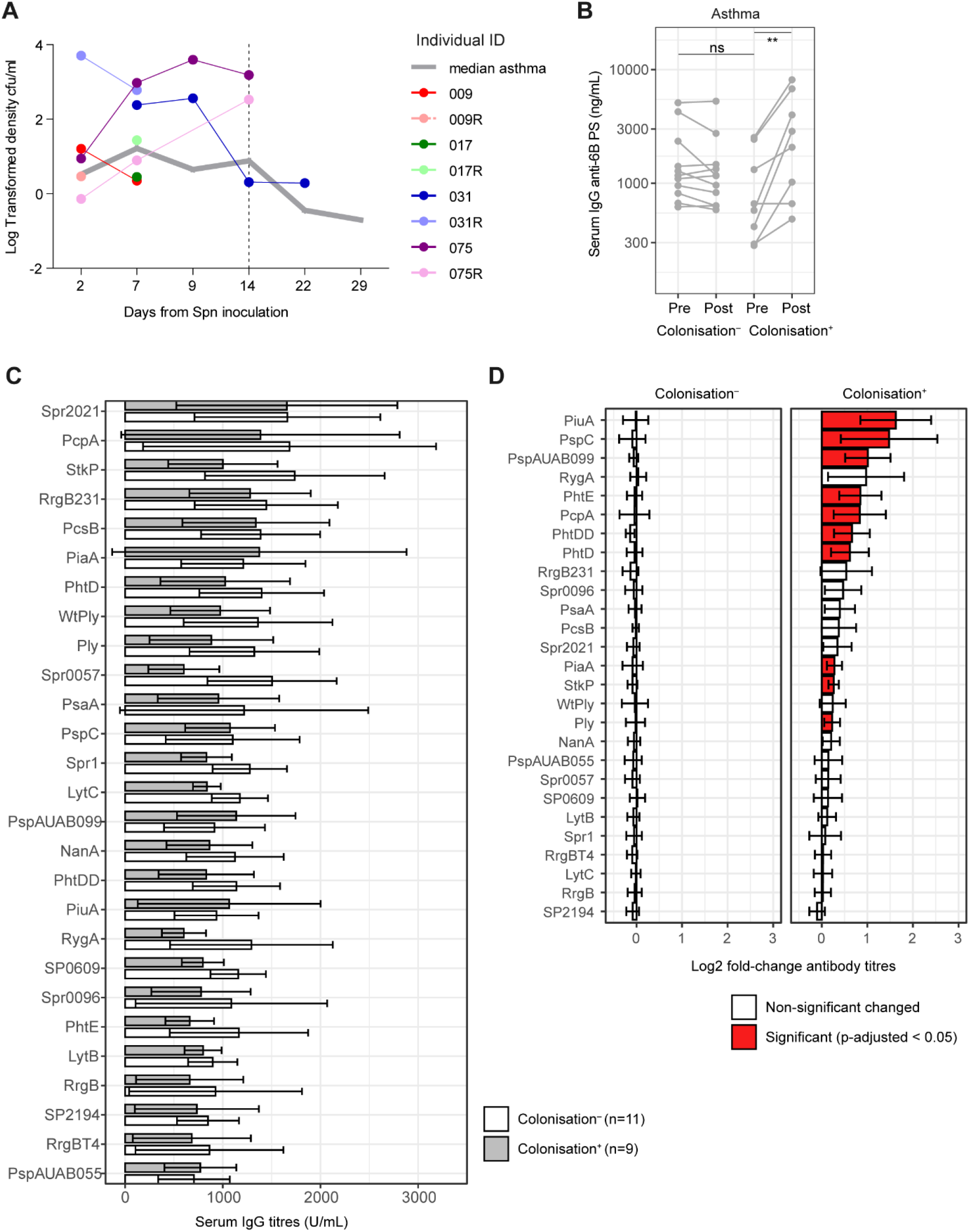
Immunising effect of colonisation in asthmatic individuals. A) Bacterial colonisation density during challenge and re-challenge. Log transformed densities at each timepoint are shown. The mean value of log-transformed colonisation density during the primary colonisation episode for all asthma individuals is depicted by a grey line (as found in Figure 1D). The four individuals that were positive upon re-challenge are depicted. Only densities for positive nasal washes are shown and re-challenge samples per volunteer are indicated by an R after their ID. For the re-challenge phase, samples were only collected up to day 14, indicated by a dashed line. B) Anti-6B polysaccharide IgG titres were measured in serum of asthmatic individuals before and after challenge, stratified by colonisation status. ** p = 0.004 by paired t-test. C) Baseline levels of IgG titres against 27 pneumococcal proteins measured by multiplex. Mean and 95% confidence intervals are shown for colonised (grey) and non-colonised (white) asthmatic individuals. D) Anti-protein IgG titres depicted as log_*2*_ fold-change to baseline, with individuals stratified by colonisation status. Mean and 95% confidence interverals are shown. Red bars depict proteins against which titres were significantly different following colonisation compared to baseline (p< 0.05 by paired t-test, corrected for multiple testing using Benjamini-Hochberg).

### Pneumococcal antibody titres before and after colonisation

In healthy controls, antibody titres against pneumococcal proteins and capsule increase following 6B experimental pneumococcal colonisation, but baseline titres do not predict of risk of subsequent acquisition. To assess this in asthma individuals, we measured IgG titres in serum against 6B polysaccharide and 27 pneumococcal proteins at baseline and day 29 post inoculation (Figure 3B–D). Anti-capsular antibody levels increased upon colonisation in asthmatic individuals (median 3-fold increase, p = 0.0039 paired t-test). However, baseline titres were not significantly different between individuals who became colonised and those who did not. Similarly, at baseline, there were no significant differences in IgG titres against any of 27 pneumococcal protein between colonised and non-colonised individuals (Figure 3C). Antibodies against 10 pneumococcal proteins were significantly induced by colonisation, including antibodies against PspC, PspA and PiuA (Figure 3D).

### Nasal cell populations in people with asthma and healthy controls before colonisation

To understand differences in immune populations underlying the altered dynamics of pneumococcal colonisation in individuals with asthma, we performed phenotyping of nasal immune cells using flow cytometry and minimally-invasive nasal curettes (Figure E3). Nasal eosinophil frequency was similar in people with well controlled asthma and healthy controls (Figure 4A and E3B), with a modest positive correlation with circulating blood eosinophil levels in people with asthma (r = 0.36, p = 0.03, Figure 4B). Nasal neutrophil frequency was lower in people with asthma compared to healthy controls (median neutrophil/epithelial cell ratio 0.35 [0.038–2.75] vs 1.72 [IQR 0.75–4.43], p = 0.0054, Figure 4C and E3C). A similar pattern was seen in CD3^+^ T cells (median T/epithelial cell ratio 0.079 [0.034–0.32] and 0.36 [0.17–0.89] in asthma and healthy controls respectively, p< 0.0001, Figure 4D and E3D). MAIT cells were specifically decreased among T cells in people with asthma (median proportion of total CD8^+^T cells 9.4% [IQR 5.0–13.3]) compared to healthy controls (median 15.8% [IQR 9.9–25.9], p = 0.0047, Figure 3E). Nasal MAIT levels correlated negatively with BMI (r = –0.38, p = 0.023, Figure 4F). Systemic MAIT cells are associated with protection against pneumococcal colonisation^22^, we analysed MAIT cells in blood of individuals with and without asthma (Figure E4). Blood MAIT cells were not different in healthy and asthma individuals (Figure 4G). MAIT cell frequency was increased at baseline in healthy individuals that were protected compared to susceptible individuals (p = 0.026); there was no difference between colonised and non-colonised asthma individuals.

**Figure 4:**
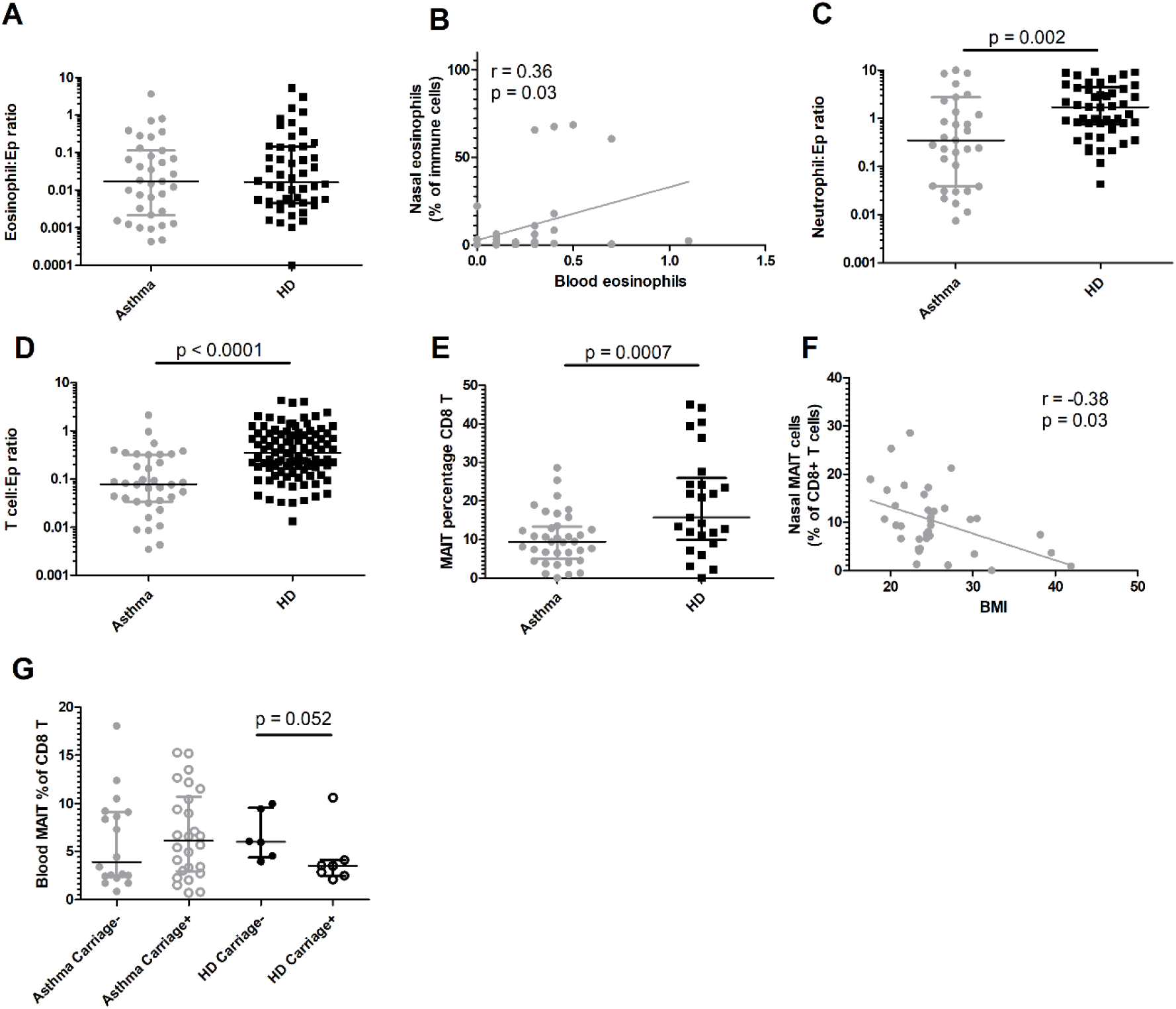
Nasal immune profiles in asthma (closed circles) and healthy control (closed squares) at baseline. Levels of A) eosinophils normalized to epithelial cells for people with asthma and healthy controls. Symbols represent individuals, with superimposed group median and IQR, and. B) Correlation by linear regression between blood eosinophils (x1000/µL blood) and nasal eosinophils (Spearman rank test). Levels of C) neutrophils and D) CD3+ T cells normalized to epithelial cells for asthmatics and healthy controls (HC). Student t-tests were performed after normalisation by log-transformation data. E) Percentage of mucosal associated invariant T cells (MAIT) amongst nasal CD8+ T cells. Median and IQR are superimposed. and symbols represent individual volunteers. Student t-test was used. F) Linear correlation between body mass index (BMI) and MAIT cell frequency. (Spearman rank test). G) Levels of MAIT cells among blood CD8+ T cells at baseline in asthmatics and healthy donors. Mann-Whitney tests were used to compare colonisation status for healthy controls or asthmatics as data were not normally-distributed.

### Changes in nasal cell populations after colonisation

There was a trend towards recruitment of monocytes at day +9 post colonisation compared to baseline in people with asthma (median fold change (FC) = 3.84, [IQR 1.15–9.38], p = 0.055). This effect of colonisation was also present in healthy controls (median FC = 3.63, [IQR 1.24–11.43], p = 0.002, Figure 5A, B). Monocyte levels remained increased at day+29 in both people with asthma (median FC = 2.23 [IQR 1.50–4.25], p = 0.03) and healthy controls (median FC = 1.72 [IQR 0.35–11.43], p = 0.049). Evidence for monocyte recruitment persisted after normalising to epithelial cells to correct for micro biopsy size (Figure 5C, D). CD66b^+^ granulocytes were significantly increased in people with asthma at day+9 (median FC = 1.78, [IQR 1.09–8.06], p = 0.044), but were no longer significantly increased at day+29 (median FC = 1.84, [IQR 0.88–5.04], p = 0.17, Figure 5E). In healthy controls, granulocytes were increased at day+7 (median FC = 3.17, [IQR 1.05–7.37], p = 0.009), returning to baseline levels at day+28 (median FC = 1.03, [IQR 0.41–3.05], p = 0.89, Figure 5F). Contraty to monocytes, In subgroup analysis, neither eosinophils or neutrophils were significantly recruited, although there was a trend for both increased neutrophils (median FC = 1.6) and eosinophils (median FC = 2.0) at day 9 in asthmatics (Figure E5). After correction to the total number of epithelial cells, these transient increases in nasal granulocyte numbers were not significant, although there was still a tendency to increased granulocyte numbers at day+9 in people with asthma (median FC = 1.90) and healthy controls (median FC = 2.07) (Figure 5G, H). No changes in monocytes and granulocytes were observed after challenge in non-colonised subjects in either asthma or healthy control cohorts.

**Figure 5.**
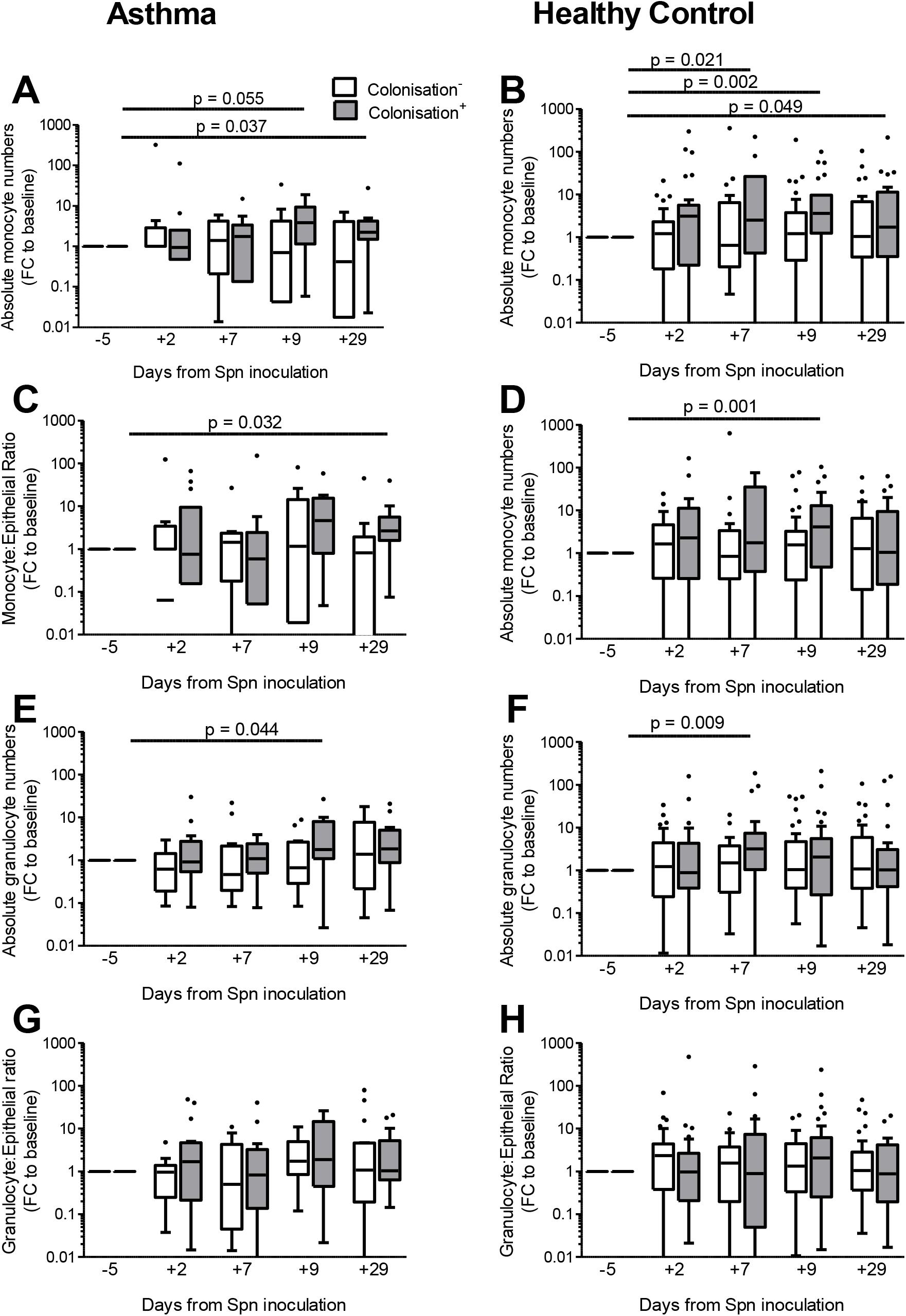
Longitudinal measurements of monocytes and neutrophils in nasal mucosa following pneumococcal challenge. Baseline-normalized levels of monocyte numbers in A) asthma and B) healthy control, and monocyte levels normalized to epithelial cells for C) asthma and D) healthy control. Baseline-normalized levels of granulocyte numbers in E) asthma and F) healthy control and granulocyte numbers normalized to epithelial cells for G) asthma and H) healthy control are shown. Tukey boxplots are presented as well as results from paired-Wilcoxon rank tests compared to baseline levels (comparing the change from baseline to the days 7, 9, and 29 as shown); FC = Fold Change, Spn = *S. pneumoniae*).

### Nasal cytokines during experimental colonisation in asthmatic and healthy individuals

We observed reduced populations of neutrophils and T cells in asthmatics at baseline (Table 2). Following colonisation, there was no evidence of excessive infiltration of inflammatory cells (Figure 4) or increase in FeNO levels (Table 2). We therefore characterized nasal inflammation in more detail by analysing nasal lining fluid for 30 cytokines and immune proteins using multiplex detection (Luminex). Recapitulating the cell population data, we saw mostly reduced cytokines in the nose of asthma compared to healthy individuals: 22/30 cytokines were significantly decreased (Figure 6A), and only anti-inflammatory IL-1RA increased. Following inoculation, we did not see any increase in nasal cytokine levels in asthma or healthy controls regardless of colonisation status (Figures 6B,C).

**Figure 6.**
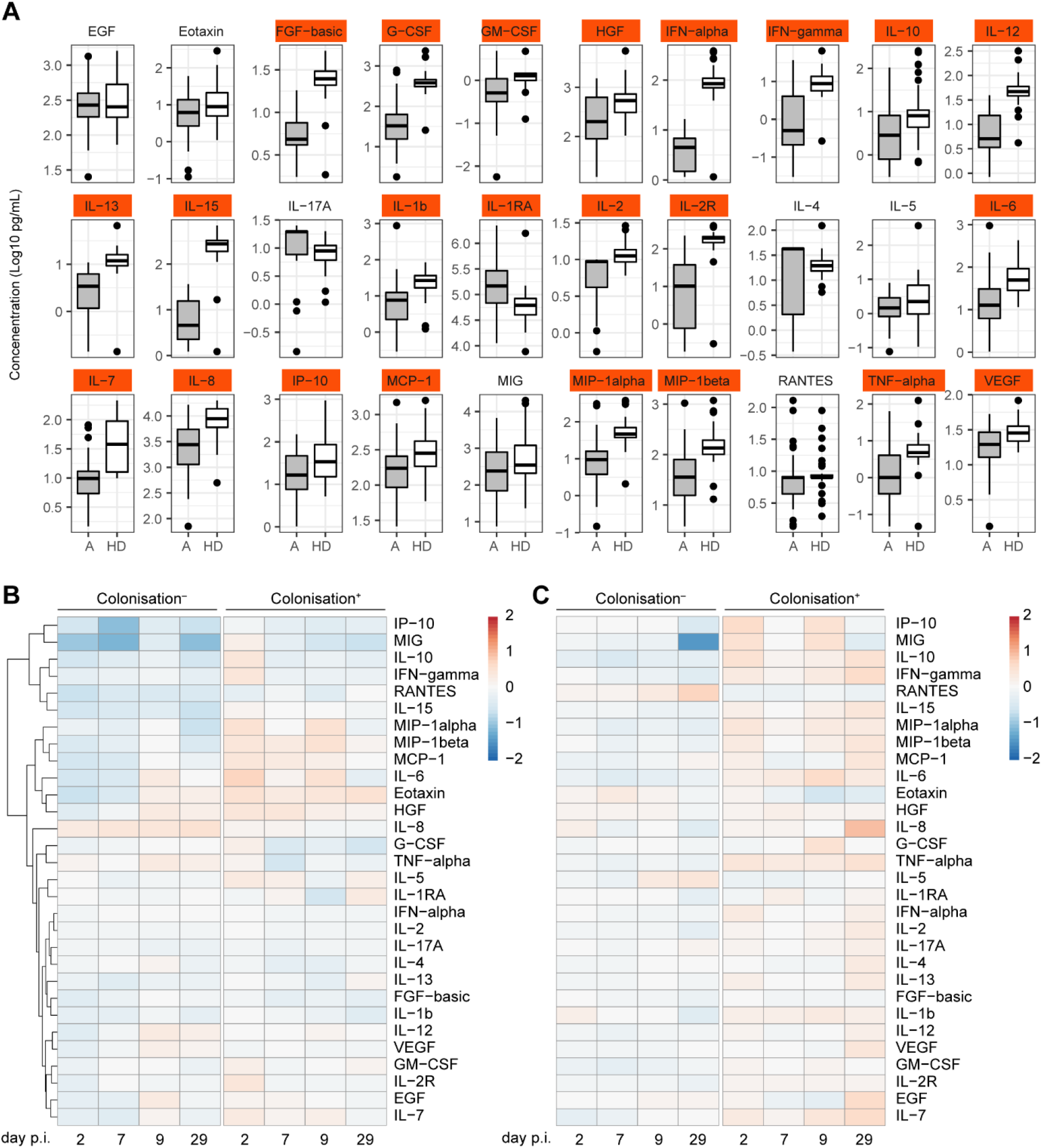
Nasal lining fluid cytokine analysis. A) Baseline levels of 30 cytokines for asthmatics (A, grey) and healthy donors (HD, white). Significantly different cytokines are indicated in red, while non-significant cytokines in white. Based on t-test on log-transformed values, followed by Benjamini-Hochberg correction for multiple testing with cut-off of p=0.01. Boxplots are shown per group. B) Asthmatic (n=55, 26 of whom were colonised) and C) healthy donor (n=63, 30 of whom were colonised) heatmaps over time following inoculation. The mean log2 fold-change compared to baseline per group is shown per day and per cytokine, stratified by colonisation status. No significant changes were seen by paired t-test followed by Benjamini-Hochberg multiple testing correction.

## Discussion

This is the first study to examine controlled challenge of an infectious bacterium in people with well controlled asthma, providing a unique opportunity to study experimental nasopharyngeal colonisation and its relation to clinical characteristics. All participants had very well controlled physician diagnosed asthma and were taking inhaled corticosteroids.

Experimental colonisation rates and density were not significantly higher in people with well controlled asthma on ICS therapy compared to healthy controls. However, a major difference to healthy comtrols is that colonisation conferred less protection to homologous re-challenge, although our sample size for re-challenge (n=12) is insufficient to determine the exact rate of protection. In previous cross-sectional observational studies, high colonisation rates in people with asthma were reported ^4^. Our observation that a colonisation episode in asthmatics is not fully immunising could explain these higher rates. However, direct translation of our results is limited as the cross-sectional data tend to have ill-defined diagnosis (self-reporting), and limited background information on medication, control, clinical characteristics and smoking history. Other factors prevalent in the wider populations of asthmatics may therefore influence colonisation rates, such as a high dose of ICS therapy, sub-optimal control, or more severe disease.

Despite incomplete protection against re-challenge, colonisation increased systemic antibody titres against pneumococcal protein and capsule. This indicates that serum IgG is not protective against colonisation in asthma, and mirrors previous data from healthy adults in which anti-capsular memory B cells gave protection, but antibody titres did not ^15^.

Nasal neutrophils and T cells, including MAIT cells, were decreased in people with asthma compared to healthy controls. Decreased MAIT cells have been reported amongst T cells in blood and sputum of people with asthma on ICS therapy ^11^. In contrast, reduced MAIT cells in this study were confined to the nasal mucosa as we did not see changes in systemic MAIT numbers in asthma compared to healthy individuals here. Studying a young cohort with well-controlled asthma may explain this.

The decreased immune frequencies of neutrophils and T cells were mirrored by decreased levels of cytokines in the nasal lining fluid of asthma individuals. Together this suggest a general immune-suppressive environment, possibly due to ICS therapy and is a reflection of the good asthma control in our participants.

After colonisation the mucosal cellular changes were similar in people with asthma and healthy controls with recruitment of monocytes and granulocytes to the nose. We have previously reported a recruitment of monocytes to the nose following colonisation in healthy volunteers ^9^; despite altered baseline nasal immune parameters, there is no impaired mucosal immune response to pneumococcal colonisation in people with asthma taking moderate doses of ICS.

We did not see significant inflammation following colonisation when measured by exhaled NO or cytokine levels. During exacerbations amongst children, pneumococcal carriage rates increase ^23^. The fact that we did not see inflammation triggered by colonisation could be an indication that pneumococcal acquisition is secondary to asthma exacerbations. Alternative explanations are that our volunteers had good asthma control, or that colonisation responses differ between children and adults. The reduced colonisation duration in asthma compared to healthy individuals was unexpected and seems paradoxical to the lack of excessive inflammation observed in asthmatic individuals. It is possible that other inflammatory or epithelial markers that we did not explore in this study could underlie this difference in ability to sustain or clear colonisation.

Within the asthma cohort, an increased BMI was associated with likelihood of experimental colonisation, while other clinical characteristics, such as FeNO, blood eosinophils and spirometry were not. Blood eosinophils and baseline FeNO levels correlated positively, as seen in previously ^24^. There was also a positive correlation between nasal and blood eosinophils. BMI has not been studied in the context of nasopharyngeal colonisation in asthma, although obesity related asthma is a recognised phenotype ^25^. Obesity may be a consequence of corticosteroid treatment, although this is unlikely in our well-controlled cohort, who required one or less course of oral steroids in the last 12 months. We do not have data on BMI in our healthy volunteers, therefore it is plausible that obesity may affect nasopharyngeal colonisation independent of asthma.

The careful characterisation of people with asthma, age-matched healthy controls and controlled challenge are a strength of our study. For safety reasons, we were unable to include participants with severe asthma on high dose of ICS. This was a small study powered to detect experimental colonisation rates in people with asthma and not general differences within the population.

## Conclusion

People with asthma had lower nasal neutrophil and T cell levels compared to healthy controls, which was confirmed by lower levels of nasal cytokines. Asthma individuals had a shorter duration of of nasopharyngeal colonisation when challenged with *S.pneumoniae*. The rate of colonisation was higher with increasing BMI, but not different from controls. Colonisation in asthma individuals had a limited immunizing effect to homologous re-challenge suggesting a function immunological difference either due to asthma disease or treatment.

## Data Availability

The datasets generated during and/or analysed during the current study are available from the corresponding author on reasonable request.

## Acknowledgements

We would like to thank all volunteers for participating in this study and the National Institute for Health Research (NIHR) Local Comprehensive Research Network for clinical support. We are also grateful to the entire EHPC team and clinicians who provided on call clinical cover for the study. Flow cytometry acquisition was performed on a BD LSR II cytometer funded by a Wellcome Trust Multi-User Equipment Grant (104936/Z/14/Z)

## Key Messages

- Duration of experimental pneumococcal colonisation is reduced in asthma
- A high BMI increases the likelihood of experimental pneumococcal colonisation in asthma
- Rate and density of pneumococcal colonisation is similar in people with asthma compared to healthy controls
- Pneumococcal colonisation does not confer protection against re-challenge in asthma, despite boosting antibody titres against pneumococcal proteins and capsule
- Mucosal immune cells and cytokine levels are reduced in asthmatic individuals with inhaled corticosteroids compared to healthy controls.
- There is recruitment of monocytes into the nasal mucosa following colonisation in people with asthma, with reduced nasal neutrophils and T cells, specifically Mucosal Associated Invariant T (MAIT) cells, in comparison to healthy controls.

## Abbreviations

ACT score: Asthma Control Test
BDP: BudesonidePropionate Equivalent
BMI: Body Mass Index
CFU: colony forming unit
EHPC: Experimental human pneumococcal challenge
FEV2: Forced expiratory volume second
FeNO: fractional Exhaled Nitric Oxide
ICS: Inhaled corticosteroids
IQR: inter quartil range
MAIT: mucosa associated invariant T cells
MSD: Meso Scale Discovery
PBMC: Peripheral blood mononuclear cells
PEFR: Peak expiratory flow rate

## Online Data Supplement

**Table E1:**
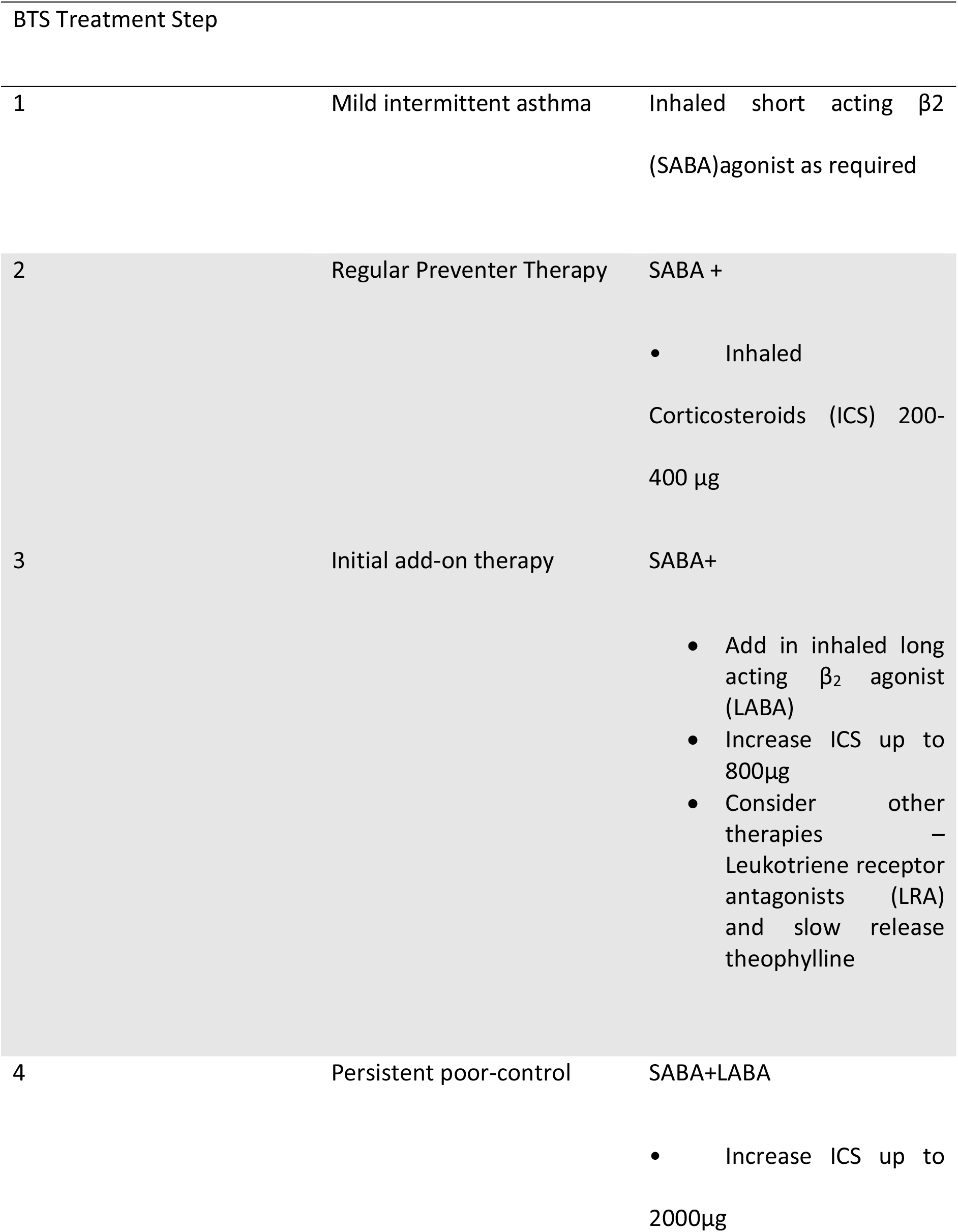
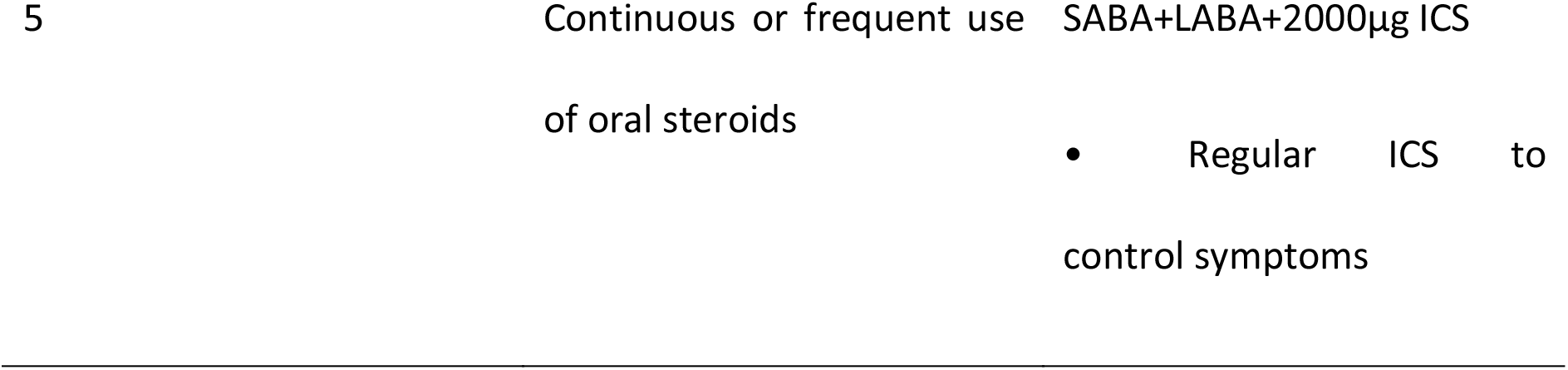
British Thoracic Society Treatment steps^26^: shaded area represent the cohort of patients recruited for the study

**Table E2:**
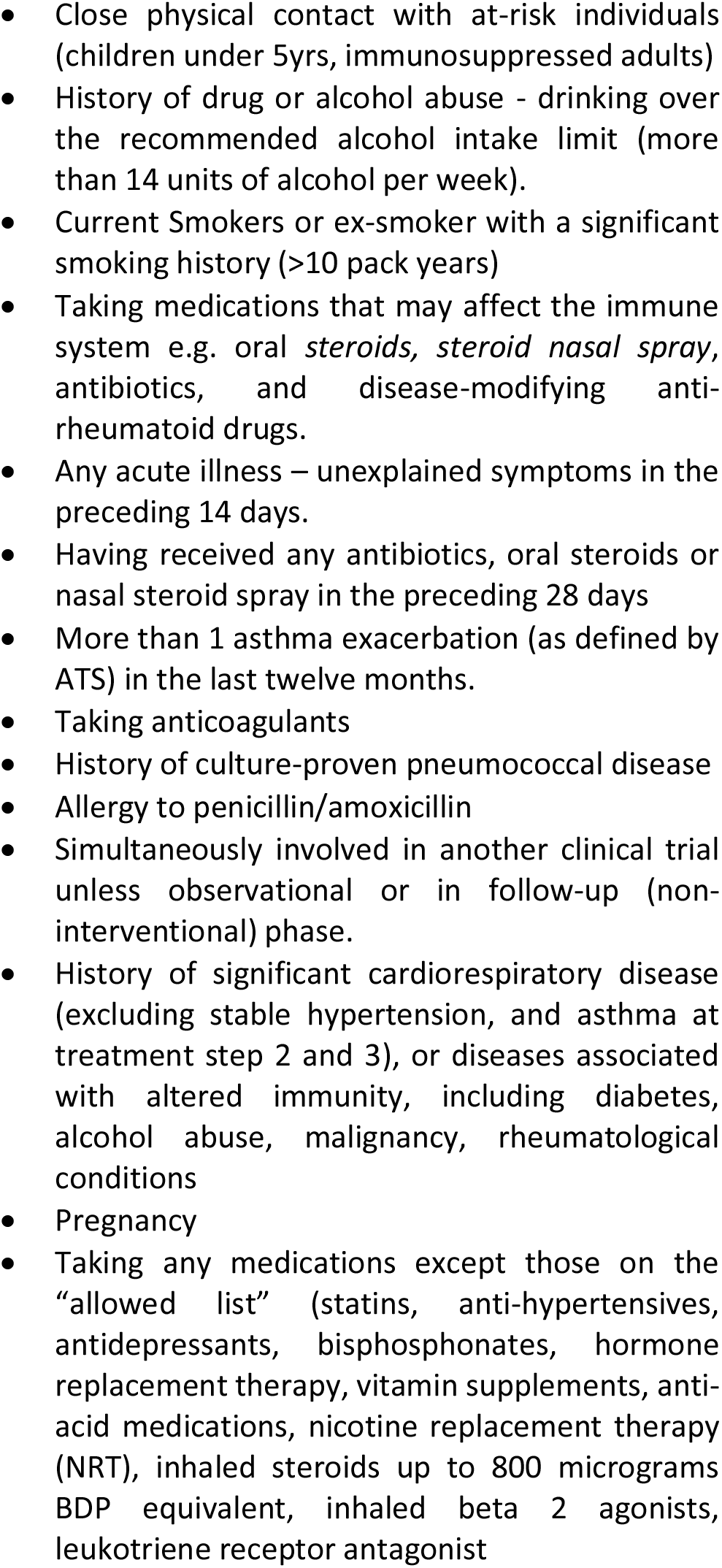
Exclusion Criteria for EHPC Asthma

**Figure E1:**
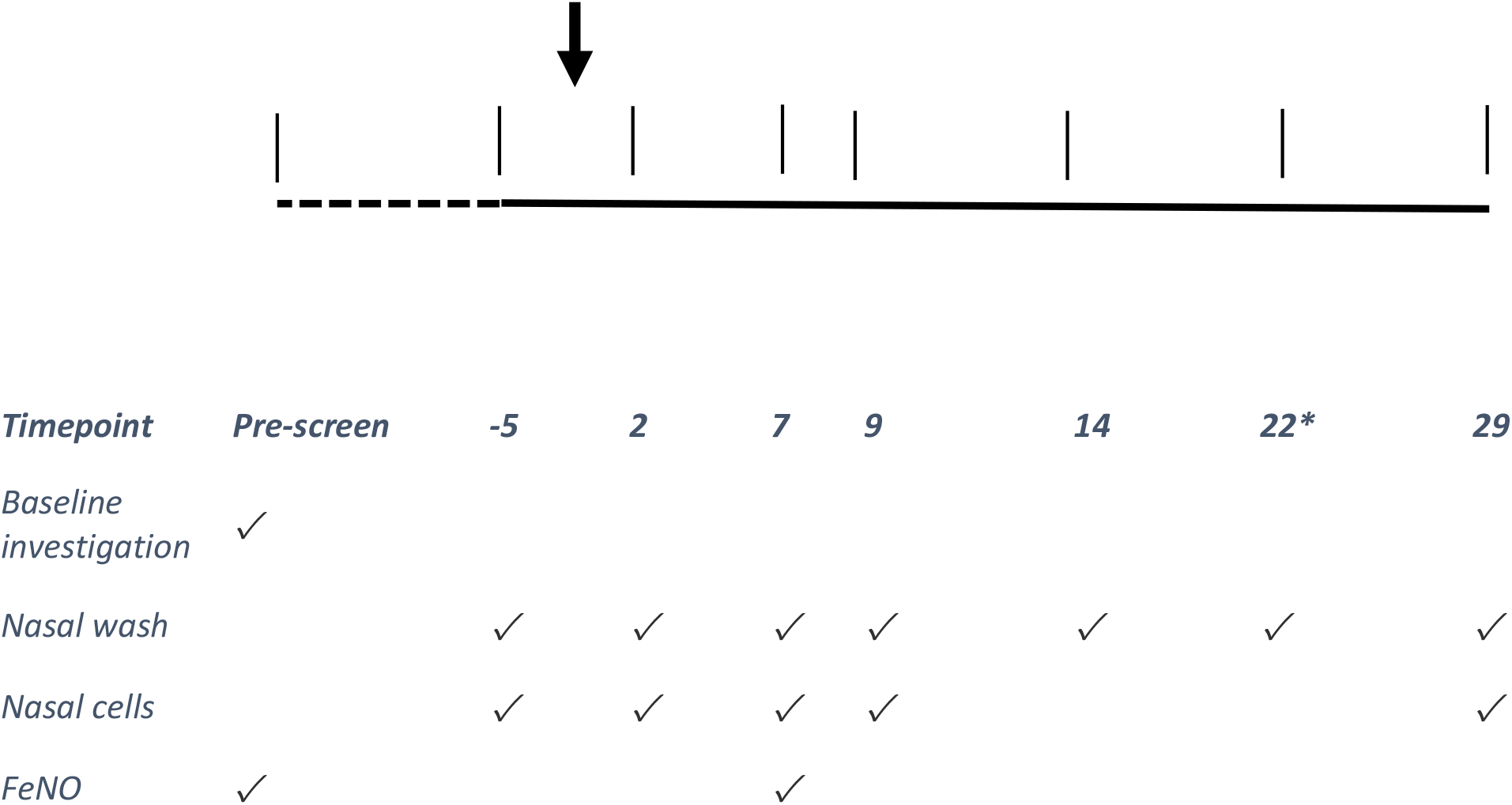
Study Overview: Study design showing timepoints for follow up visits. Participants were pre-screened up to 14 days before screen; at baseline full history, including relevant examination was done along with Spirometry, Peak Expiratory Flow Rate (PEFR), and baseline blood samples. Nasal wash was obtained for pneumococcal detection and nasal cells for immunophenotyping. *Only colonised participants. Fractional Exhaled Nitric Oxide (FeNO)

**Figure E2:**
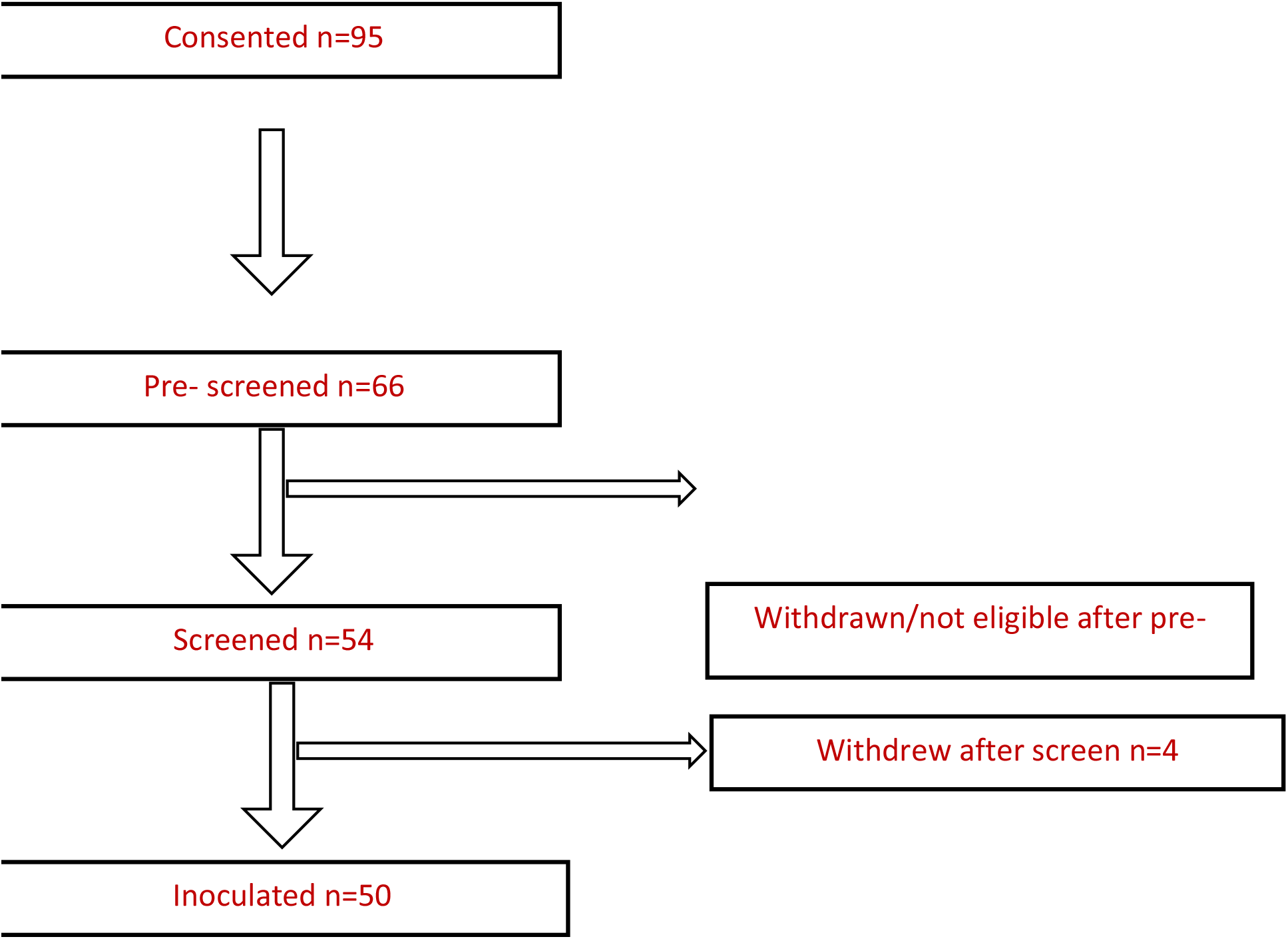
Recruitment: Flow chart showing recruitment and retention numbers. Reasons for withdrawal: Not on inhaled corticosteroids (ICS) n = 17, unable to contact after consent n = 5, medical placement n = 2, ICS dose too high n = 2, n = 1 for each of the following: not using ICS as prescribed, moved out of the area, no reason stated, personal reasons, high white cell count, headaches daily, penicillin allergy, bronchiectasis, using non-invasive ventilation, withdrew consent, > 3 exacerbations in the preceding 12 months, proven pneumococcal pneumonia. One participant was excluded after day 2 as they had low peak expiratory flow rate (PEFR) and had to increase ICS, thus exceeding the 800µg budesonide equivalent threshold.

**Figure E3:**
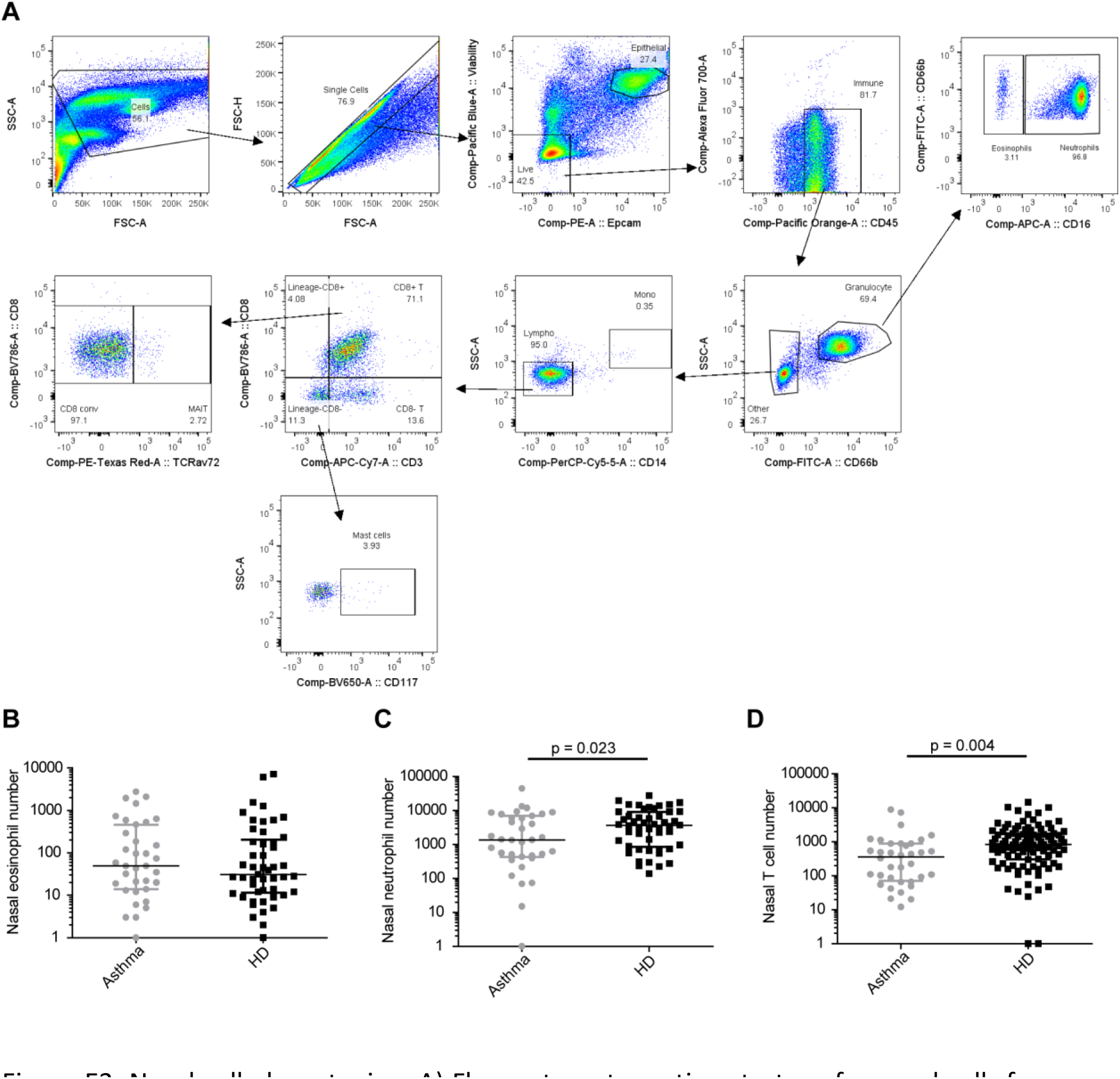
Nasal cell phenotyping. A) Flow cytometry gating strategy for nasal cells from one representative participant with asthma. Numbers of acquired B) eosinophils, C) neutrophils and D) T cells from the nose at baseline for asthmatic (grey) and healthy (black) donors. Median and interquartile ranges are depicted, as well as results from student t-test on log-transformed data.

**Figure E4:**
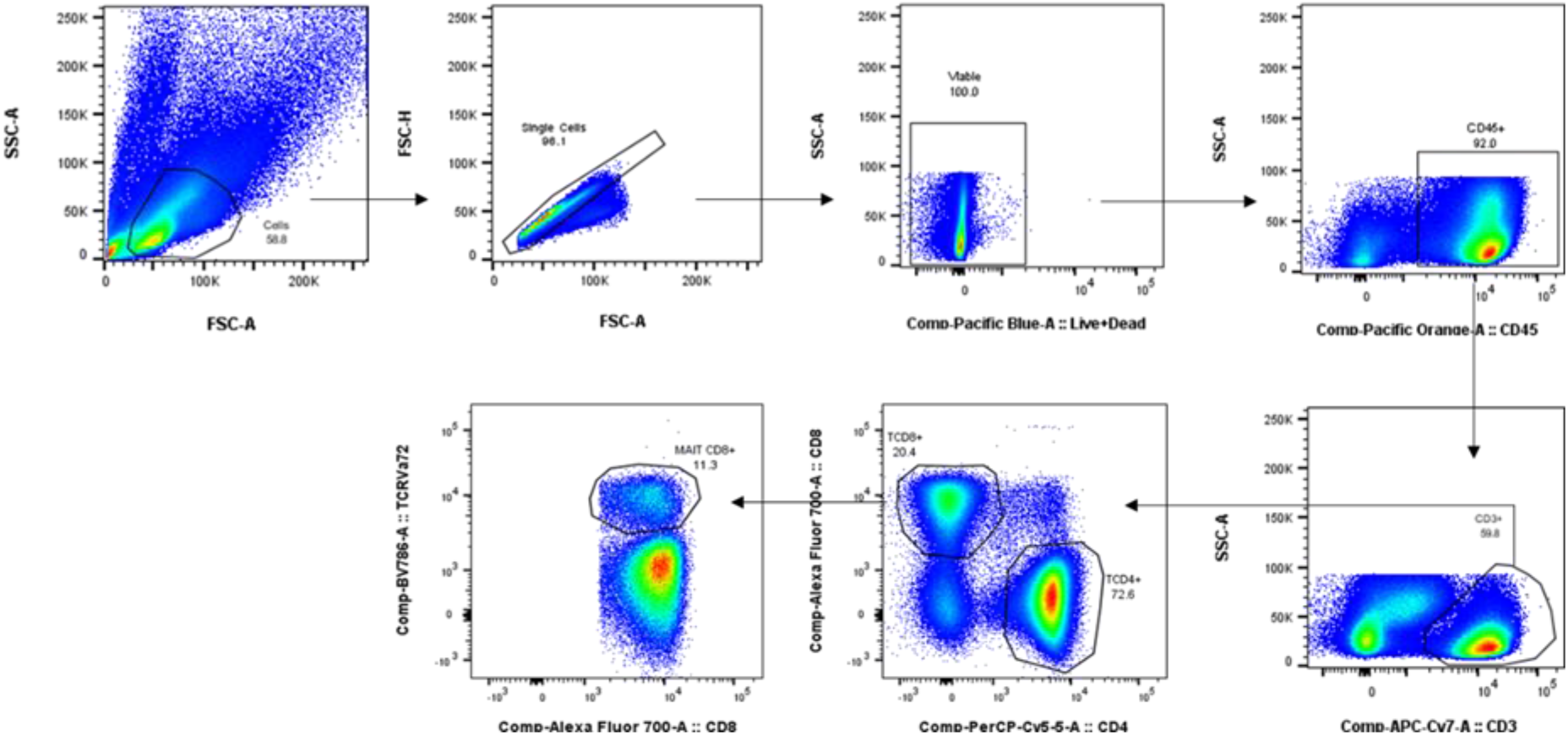
Blood MAIT cell phenotyping. Flow cytometry gating strategy for blood cells from one representative participant with asthma.

**Figure E5.**
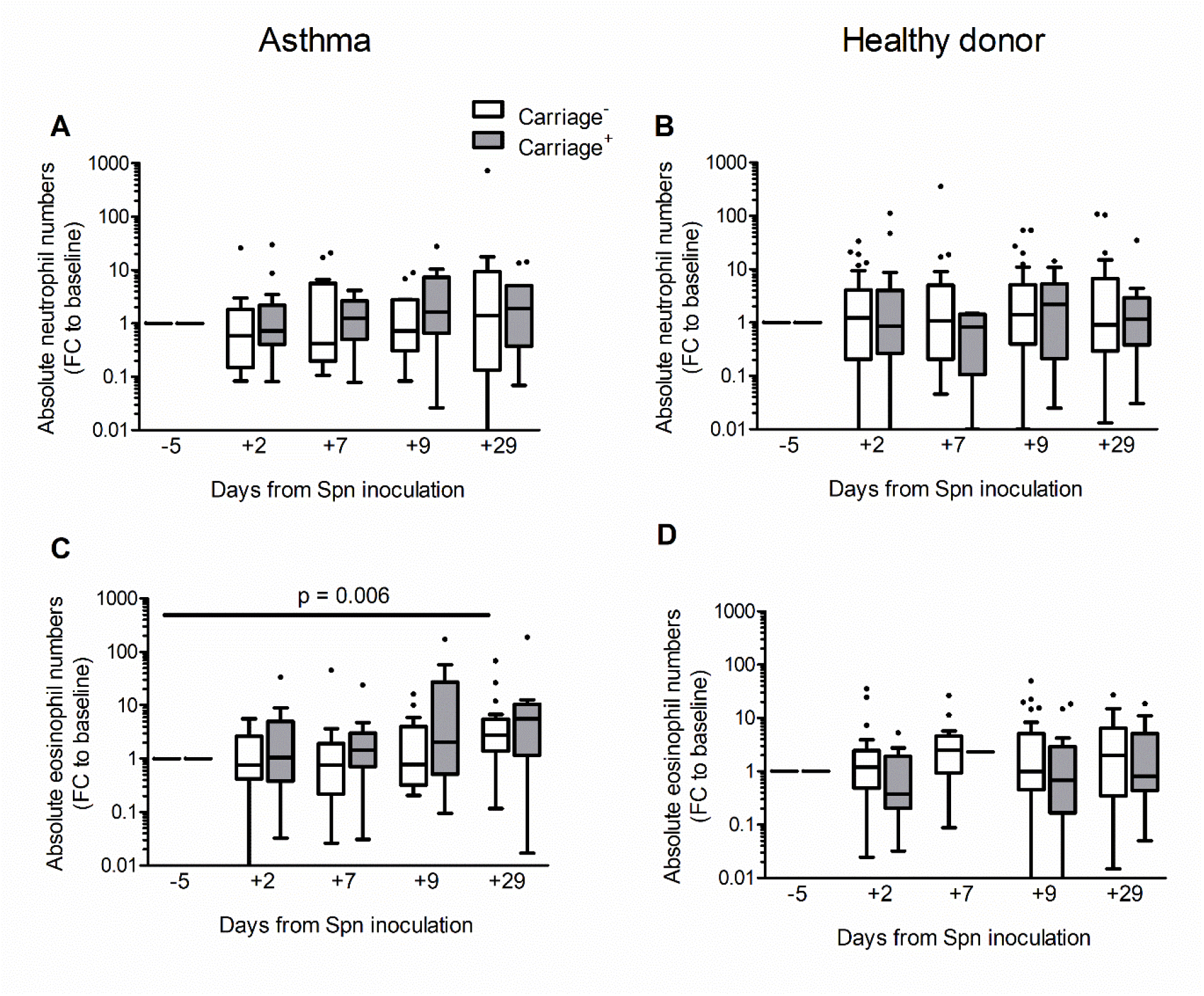
Nasal neutrophil and eosinophil numbers. Baseline-normalized numbers of neutrophils in A) asthma and B) healthy control. Baseline-normalized numbers of eosinophils in C) asthma and D) healthy control. Tukey boxplots are presented as well as results from paired-Wilcoxon rank tests compared to baseline levels (comparing the change from baseline to the days 7, 9, and 29 as shown); FC = Fold Change, Spn = *S. pneumoniae*).

## Notes

**Funding:** The project was funded by MRC programme grant (MR/M011569/1) to SG, and Bill and Melinda Gates Foundation (grant OPP1117728). Flow cytometric acquisition was performed on a BD LSR II cytometer funded by a Wellcome Trust Multi-User Equipment Grant (104936/Z/14/Z).

**Conflict of Interest:** The authors of this study do not have any disclosures or conflict of interests to declare

### Competing Interest Statement

The authors have declared no competing interest.

### Clinical Trial

International Randomised Controlled Trial Number (ISRCTN) 16755478

### Funding Statement

The project was funded by MRC programme grant (MR/M011569/1) to SG, and Bill and Melinda Gates Foundation (grant OPP1117728). Flow cytometric acquisition was performed on a BD LSR II cytometer funded by a Wellcome Trust Multi-User Equipment Grant (104936/Z/14/Z).

### Author Declarations

Ethical Approval was obtained from the Liverpool East NHS Research Ethics Committee (reference number NW/016/0124). The study was co-sponsored by the Royal Liverpool University Hospital and the Liverpool School of Tropical Medicine. International Randomised Controlled Trial Number (ISRCTN) 16755478). Control group was obtained from the study approved under Liverpool East NHS Research Ethics Committee (EudraCT 2014-004634-26)

